# Mechanically Inducing Gait Abnormalities to Evaluate the Equivalence of the StrideLink Gait Device to Motion Capture

**DOI:** 10.64898/2026.01.23.26344735

**Authors:** Aaron Henry, Carson Benner, Cassandra McIltrot, Andrew B. Robbins

**Author notes:** Corresponding author: Andrew Robbins.

## Abstract

**Background:** Inertial measurement units (IMUs) have potential to be inexpensive, portable sensors for collecting gait parameters and joint kinematics. Current validation protocols generally do not investigate IMU accuracy in measuring altered gait; therefore, they cannot assess an IMU’s ability to detect pathologies. The Stridelink IMU-based gait analysis device is intended for use in detecting and monitoring gait abnormalities, thus there is a need to evaluate the device’s accuracy under abnormal gait conditions.

**Research question:** How well do measurements from the StrideLink IMU agree with motion capture (MoCap), particularly when gait is mechanically altered to simulate pathology?

**Methods:** Twenty-eight healthy participants (ages 18-40) were analyzed during a one-minute treadmill walk with Vicon MoCap and StrideLink. Tests were performed under normal and mechanically induced abnormal conditions (knee brace, walking boot). Equivalence testing and correlation analysis evaluated StrideLink’s accuracy against MoCap.

**Results:** StrideLink showed statistical equivalence (within 5%) for average cadence, stride, swing, and stance times but not double support time. Many metrics were statistically equivalent (p < .001) despite induced abnormalities. Correlation analysis showed almost perfect agreement with MoCap for stride times, cadence, and stance. However, the abnormal gait protocol revealed nuances not observed in normal gait; specifically, the device underestimated swing time by ∼10 ms in knee brace restricted limbs.

**Significance:** This study utilized mechanically induced gait abnormalities to assess the robustness of IMU measurements. Results indicate StrideLink yields valid temporal gait measurements comparable to reference systems, even under conditions of significant deviation, supporting the utility of using induced abnormalities for sensor validation.

## 1. Introduction

A healthy human gait cycle is inherently cyclical and symmetrical. Gait pathologies cause alterations to this cycle that are primary symptoms of musculoskeletal and neurological disorders. Although qualitative methods such as patient self-reporting and clinician observation are useful, quantitative analysis methods are required for accurate determination of spatiotemporal metrics of gait, which can provide insight into disease processes. By utilizing inertial measurement unit (IMU) technology, these changes can be measured outside of a controlled clinical setting, allowing for long-term and regular measurements of an individual’s everyday walking abilities. Indicators of walking instability include changes in left to right gait phase durations and double support time, while measurements such as stride time and cadence provide insight into changes in pace. The clinical significance of these metrics has been investigated across orthopedic and neurological care, highlighting the potential for at-home measurement of gait parameters to be meaningful clinical metrics of disease progression and/or healing.[1–6].

Among the various methods for quantitative gait analysis, marker-based optical motion capture systems are often considered the “gold standard” and are routinely used in clinical studies of gait and motion [7–14]. However, these systems are resource intensive, requiring complex setup, expensive equipment, and dedicated lab or clinical space for use, making them unsuitable for at-home patient monitoring [1, 7, 9, 14]. Within the past 20 years, devices utilizing wearable inertial measurement units (IMUs) have emerged as promising systems for gait analysis capable of high accuracy and reliability [2, 15–17]. An IMU typically consists of a set of gyroscopes, accelerometers, and magnetometers inside a small housing [10]. Systems utilizing IMUs as the primary method for data acquisition offer an inexpensive, portable, and relatively simple set up for data collection. When worn by a user, an IMU can measure normal and abnormal gait kinematics, which clinicians can analyze for abnormalities indicative of specific gait pathologies [1, 14, 18]. However, IMUs still present several limitations, including complex calibration requirements, drift effects that reduce accuracy over time, and limitations to wearability. Taken together, these concerns have limited the clinical utility of currently available tools [6, 19, 20].

The primary purpose of this work is to investigate the equivalence of a new commercial IMU based device (StrideLink Inc.), that aims to address many of the limitations of alternative systems, to precise measurements from an optical motion capture system (Vicon Inc.). However, because the purpose of the StrideLink device is to monitor changes and abnormalities in gait, it was deemed important that the device be tested under both normal and abnormal gait conditions. Current IMU validation techniques rarely explore abnormal gait patterns [13, 21, 22], and only a few studies have attempted to simulate gait deviations for IMU gait data analysis [14, 23–26]. Therefore, in furtherance of the goal of evaluating the StrideLink device under abnormal gait, this work also describes a novel method for producing abnormal gait that may have utility in future validation studies of similar devices.

## 2. Methods

### 2.1. Participants

31 healthy persons between the ages of 18 and 40 (mean age: 27.16 ± 4.59 years old) were recruited to participate in this study. Both males (n = 10, mean age: 26.70 ± 3.53 years old) and females (n = 21, mean age: 27.38 ± 5.08 years old) participated in this study. Participants voluntarily enrolled in the study and were excluded if they had any major orthopedic injuries preventing them from walking on a treadmill, walking on a treadmill with an ankle boot, or walking on a treadmill with a knee brace. Two participants were removed due to potential memory corruption of the StrideLink device during the trials, and one subject was removed due to wearing attire that was not compatible with the motion capture markers. Consequently, 28 subjects were included in the final analysis.

### 2.2. Data Collection

Each subject was fitted with retroreflective markers following the guidelines for the Vicon lower body Plug-In Gait model (Figure 2), and the StrideLink device was clipped to the dorsum of each shoe (over the laces) as shown in Figure 3. Calibration is not required by the StrideLink sensors due to the unique methods of data processing (described further in Section 2.3). A motion capture system consisting of 12 optical cameras (Vantage-V16, Vicon Inc., Oxford, United Kingdom) was used to track the participant’s motion while they performed a series of walking trials on a treadmill (Sunny Health & Fitness Walking Treadmill, SF-T7857) (Figure 1). Each participant completed 5 trial types; 1) normal gait, 2) constrained right ankle, 3) constrained left ankle, 4) constrained right knee, and 5) constrained left knee. Participants performed each trial type up to 3 times to ensure a sufficient number of gait cycles was collected. Each trial included a slow increase (ramp up) treadmill speed, at least 1 minute of steady state walking at the participant’s chosen speed, and a slow decrease (ramp down) in speed.

**Figure 1:**
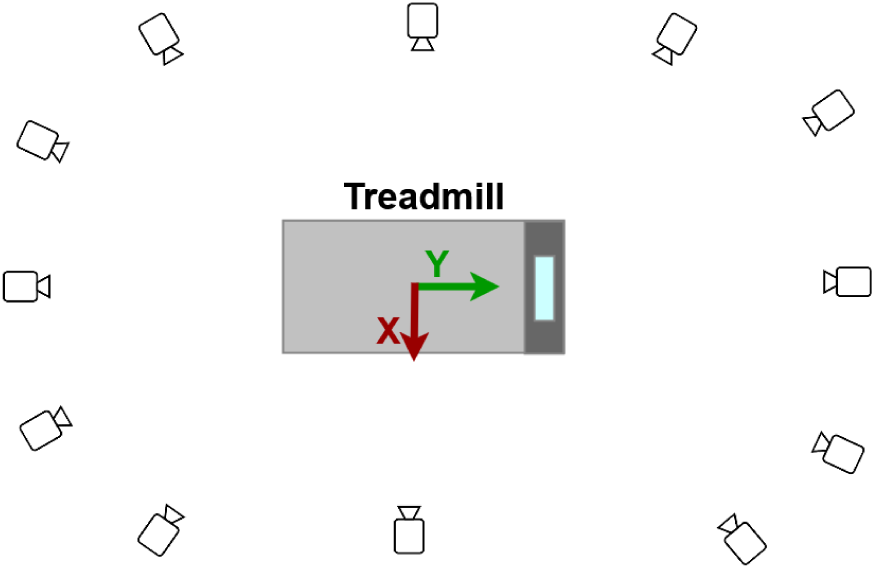
Vicon motion capture system camera layout and treadmill.

**Figure 2:**
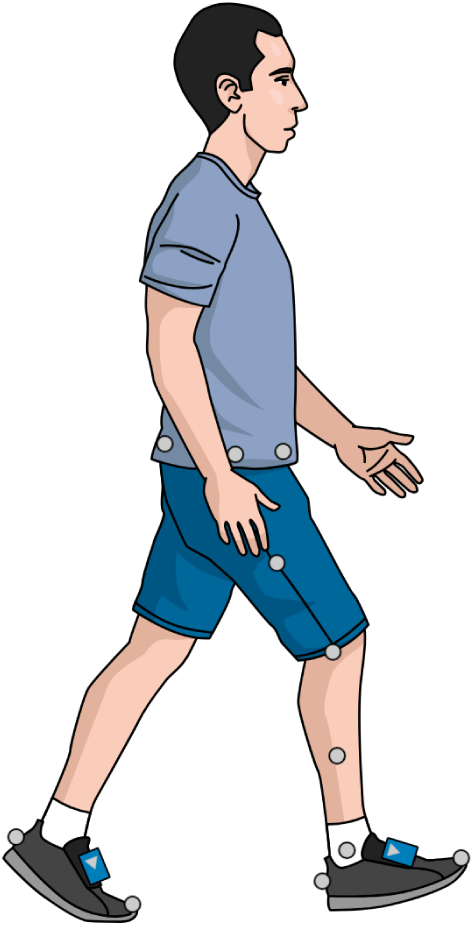
Vicon Plug-In Gait lower body marker set and StrideLink devices.

**Figure 3:**
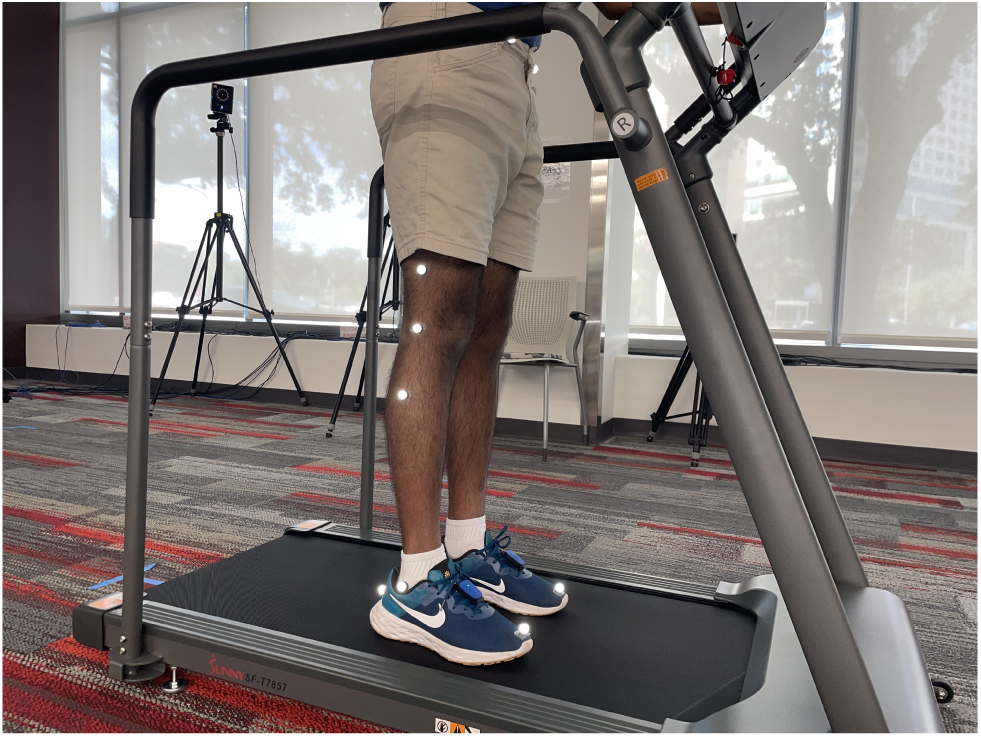
Photograph of marker and IMU placement.

#### 2.2.1. Simulating Abnormal Gait

To induce abnormal gait, two devices were used; 1) an ankle boot (United Ortho Short Air Cam Walker Fracture Boot, USA14113) on either the left or the right ankle that restricted mobility of the ankle, and 2) a knee brace (Vive ROM knee brace, SUP1088) worn on either the left or right knee. The ankle boot completely restricted ankle and foot motion, whereas the knee brace did not completely restrict motion, but rather restricted flexion/extension of the knee to 30 degrees of flexion and greater (preventing extension beyond 30^◦^ of flexion).

### 2.3. Data Processing

#### 2.3.1. Processing of Motion Capture Measurements

Data processing and analysis was conducted in Python 3.9.12. Custom scripts were used for event detection, gait metric calculations, and statistical analysis. The ramp up and ramp down phases of each trial were cropped out to include only the steady state portion of the trial in motion capture analysis.

Prior to event detection, motion capture marker trajectories were filtered with a lowpass 4th order Butterworth filter with a 4Hz cutoff frequency. Event detection was performed using the velocity-based treadmill algorithm from Zeni *et al.* [27]. The point when the horizontal component of the toe marker velocity changed from negative to positive was used to determine terminal contact (TC). The point when the horizontal component of the heel marker velocity changed from positive to negative was used to determine initial contact (IC). Event detection for motion capture data was limited to the steady state section of treadmill walking, and the steady state portion of the trial was cropped from the first terminal contact to the last terminal contact.

The gait metrics calculated were stride time, cadence, double support time, swing time, and stance time, in alignment with the metrics calculated by the StrideLink Device. Notably, the StrideLink device does not generate certain spatial measurements such as step height and step length, so those metrics were not calculated from the motion capture measurements either. Stride time was calculated as the time between IC and next consecutive IC for each foot; cadence was calculated as the number of steps per minute of steady state walking, where a step was defined as the time between IC for one foot and IC for the contralateral foot; double support time was calculated as the average time in which both feet were on the treadmill for every gait cycle; swing time was calculated as the difference between TC and next consecutive IC for each foot; and stance time was calculated as the difference between IC and next consecutive TC for each foot. Gait metrics were calculated for each subject and trial type (normal gait, ankle boot, and knee brace), and in the case where a single subject repeated a trial type multiple times, multiple trials were combined into a single dataset for each subject.

#### 2.3.2. StrideLink Device Processing

The StrideLink device functions automatically, and no user input was provided during the experiment to alter the processing results of the devices. Internally, the StrideLink device identified suitable gait cycles to analyze based on a proprietary algorithm which detects gait events using maxima and minima values of the acceleration magnitude. Only steps that were paired between the left and right sensors were used in order to classify steady state gait, and toe off, heel strike, and midswing events were identified in order to calculate temporal metrics. The gait metrics calculated by the device include stride time, cadence, double support time, swing time, and stance time. Absolute error was calculated as the difference between motion capture and StrideLink measurements while relative error was the absolute error expressed as a percentage of the motion capture measurement.

### 2.4. Statistical Analysis

Paired equivalence tests were used to determine if a metrics calculated from the motion capture data and the StrideLink device data were statistically equivalent. Equivalence bounds were set at 5% of the mean value calculated from the motion capture data. Equivalence tests used mean values from each subject to determine an overall equivalence across all participants. Equivalence tests were performed in Python using the Statsmodel module version 0.13.2.

Correlation analysis was also performed using both Pearson’s correlation coefficients (PCC) and Lin’s correlation coefficients (LCC). Interpretation and thresholds of correlation coefficients were taken from Akoglu *et al.* [28].

## 3. Results

Stride time was statistically equivalent (p < 0.001) between motion capture and the StrideLink device across all trial types and feet (Figure 4). Both left and right stride time measured with the StrideLink device were very highly correlated (PCC > 0.9) and had almost perfect agreement (LCC > 0.99) with motion capture measurements (Supplemental Tables 1-2). Stride time absolute error was consistently negative at approximately -3.75ms on average showing that the StrideLink device slightly underestimated stride time.

**Figure 4:**
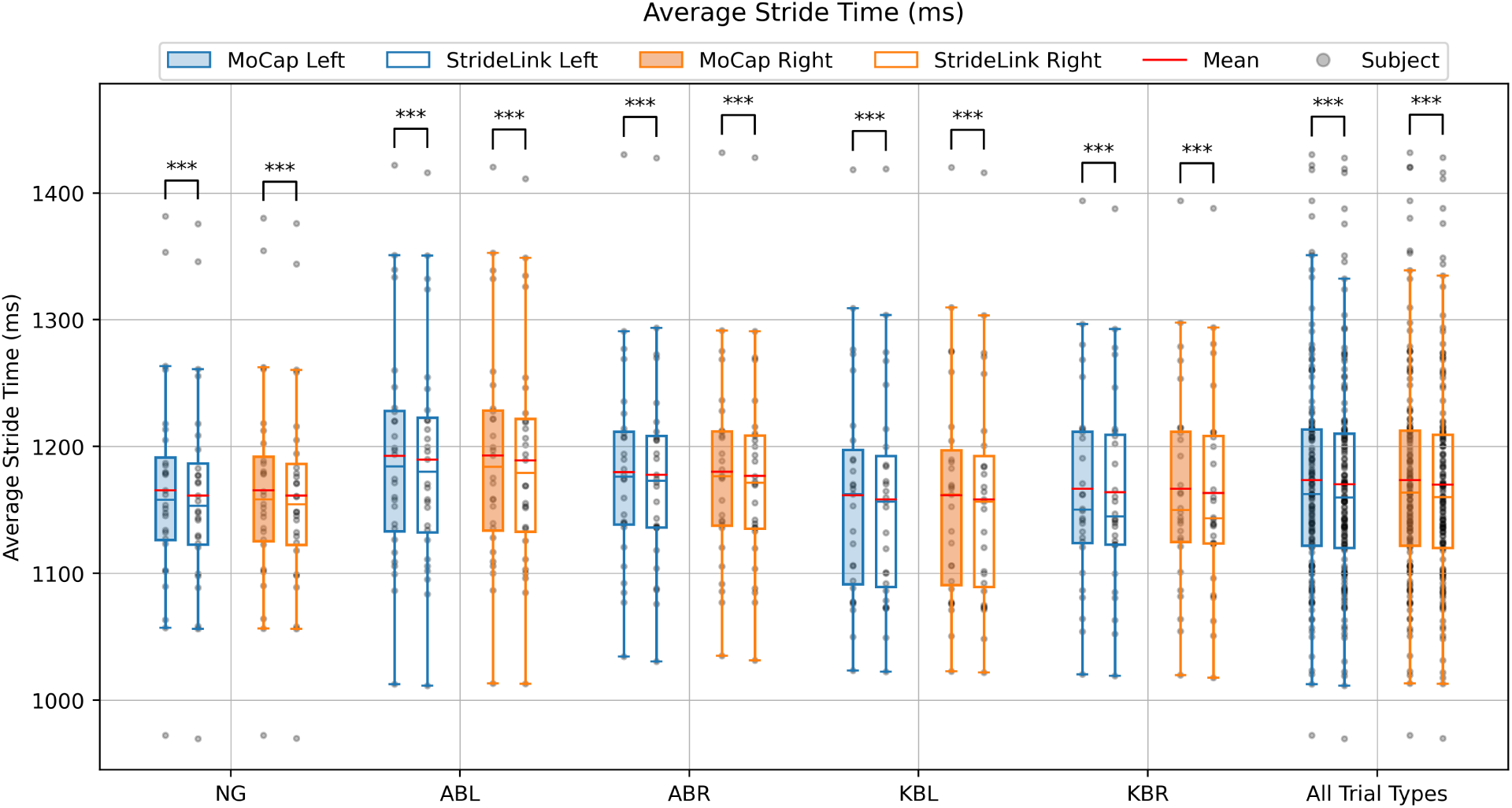
Boxplots of average stride time measured with motion capture and the StrideLink device. Motion capture boxplots are shaded and device boxplots are not. Left lower limb metrics are shown in blue, and right lower limb metrics are shown in orange. Mean values are shown as a red line within each boxplot. Individual subject mean data points are shown as points. Trial type abbreviations are as follows: NG - normal gait, ABL - ankle boot left, ABR - ankle boot right, KBL - knee brace left, KBR - knee brace right.

Average cadence measurements were significantly equivalent (p < 0.001) between the StrideLink device and motion capture data (Figure 5). Cadence values measured with the device were very highly correlated (PCC > 0.9) and had almost perfect agreement (LCC > 0.99) with motion capture measurements (Supplemental Tables 1-2). Average cadence relative errors showed that the StrideLink device consistently overestimated cadence by approximately 1% of the motion capture measurements.

**Figure 5:**
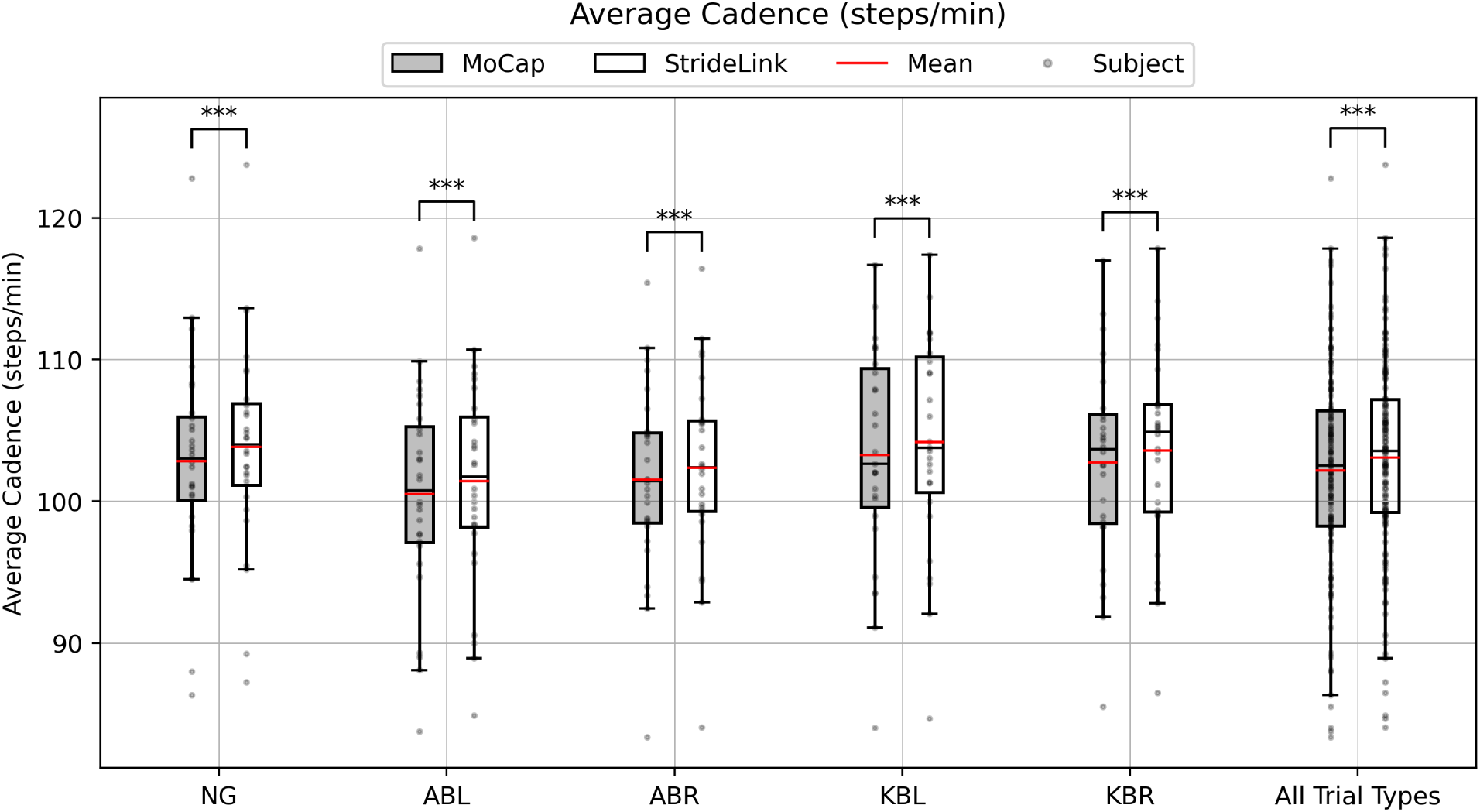
Boxplots of average cadence measured with motion capture and the StrideLink device. Motion capture boxplots are shaded and device boxplots are not. Left lower limb metrics are shown in blue, and right lower limb metrics are shown in orange. Mean values are shown as a red line within each boxplot. Individual subject mean data points are shown as points. Trial type abbreviations are as follows: NG - normal gait, ABL - ankle boot left, ABR - ankle boot right, KBL - knee brace left, KBR - knee brace right.

Double support time was significantly equivalent (p < 0.001) between the two measurement methods for trials with an ankle boot. Measurements were significantly equivalent (p < 0.05) when all trials were taken together. However, measurements were not significantly equivalent (p > 0.05) for normal gait trials and trials with a knee brace (Figure 6). All trial types were highly correlated (PCC > 0.9) between motion capture and device data. Only normal gait and ankle boot trials had moderate agreement (0.9 < LCC < 0.95) between motion capture and device measurements (Supplemental Tables 1-2). Average double support percentages followed similar trends to average double support time in both equivalence tests. Correlations for double support percentage were slightly lower than average double support time correlations.

**Figure 6:**
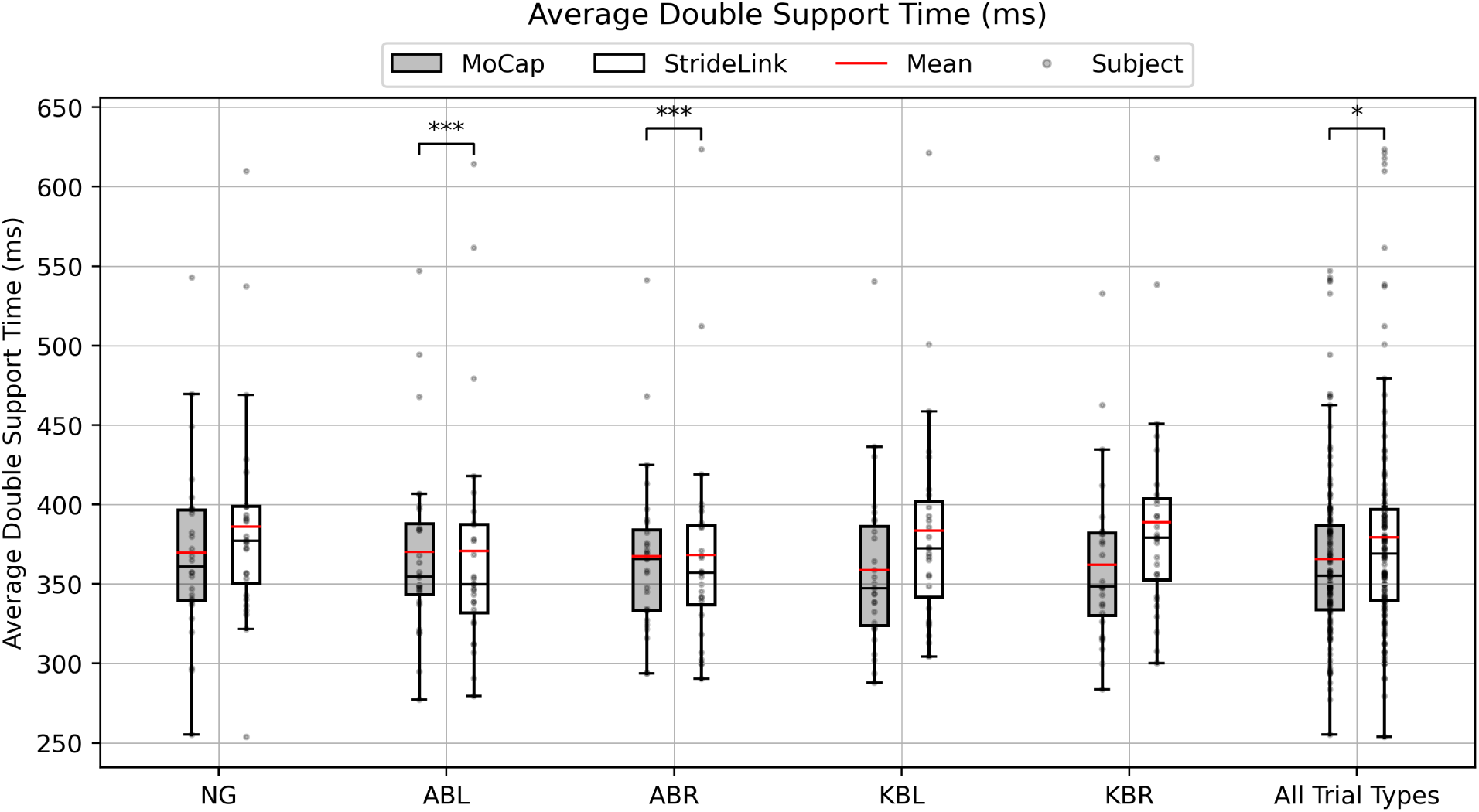
Boxplots of average double support time measured with motion capture and the StrideLink device. Motion capture boxplots are shaded and device boxplots are not. Left lower limb metrics are shown in blue, and right lower limb metrics are shown in orange. Mean values are shown as a red line within each boxplot. Individual subject mean data points are shown as points. Trial type abbreviations are as follows: NG - normal gait, ABL - ankle boot left, ABR - ankle boot right, KBL - knee brace left, KBR - knee brace right.

Average swing time was significantly equivalent (p < 0.001) across all trial types and feet except on the foot that the knee brace was on. In this case, the measurements were still equivalent at p < 0.05 (Figure 7). Average swing time of the left foot was high (PCC > 0.7) to very highly (PCC > 0.9) correlated between the measurement methods, but the measurements had poor agreement (LCC < 0.9). Average swing time of the right foot was high (PCC > 0.7) to very highly (PCC > 0.9) correlated between the measurement methods, but the measurements had poor (LCC < 0.9) to moderate agreement (0.9 < LCC < 0.95) (Supplemental Tables 1-2). Swing time as measured by the StrideLink device was generally underestimated for both feet across all trials. Notably, the device appeared to increase in error when the device was attached to the ankle boot. Average swing percentages followed similar trends to average swing time in both equivalence tests and correlation analysis.

**Figure 7:**
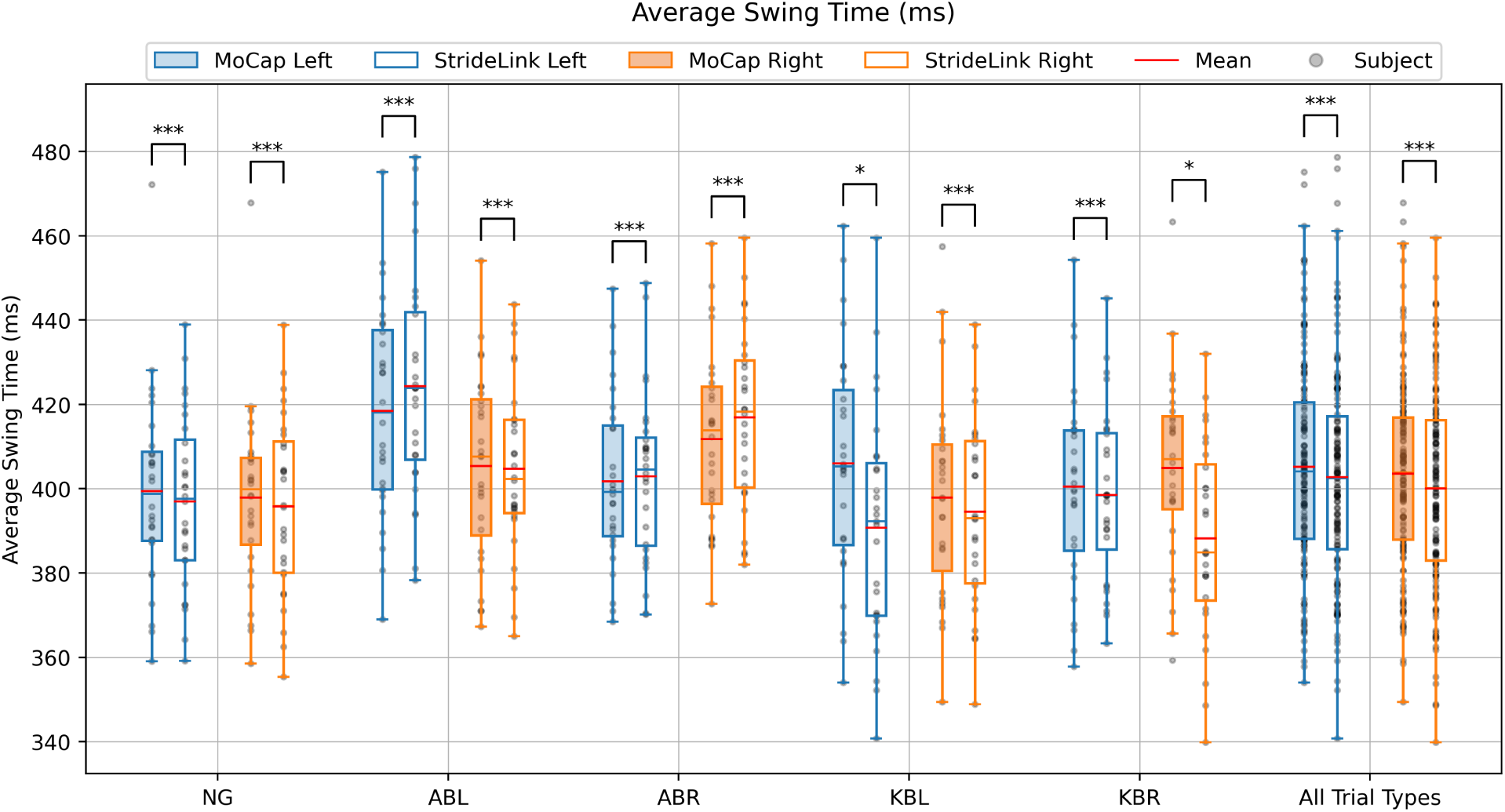
Boxplots of average swing time measured with motion capture and the StrideLink device. Motion capture boxplots are shaded and device boxplots are not. Left lower limb metrics are shown in blue, and right lower limb metrics are shown in orange. Mean values are shown as a red line within each boxplot. Individual subject mean data points are shown as points. Trial type abbreviations are as follows: NG - normal gait, ABL - ankle boot left, ABR - ankle boot right, KBL - knee brace left, KBR - knee brace right.

Average stance time was equivalent across all trial types and feet (p < 0.001) (Figure 8). Average stance time was highly correlated (PCC > 0.9) and had substantial agreement (0.95 < LCC < 0.99) between motion capture and StrideLink device measurements across all trial types (Supplemental Tables 1-2). Average stance percentages were equivalent across all trial types and feet (p < 0.001), however, these percentages had lower Pearson correlation coefficients and generally had poor agreement between motion capture and device measurements.

**Figure 8:**
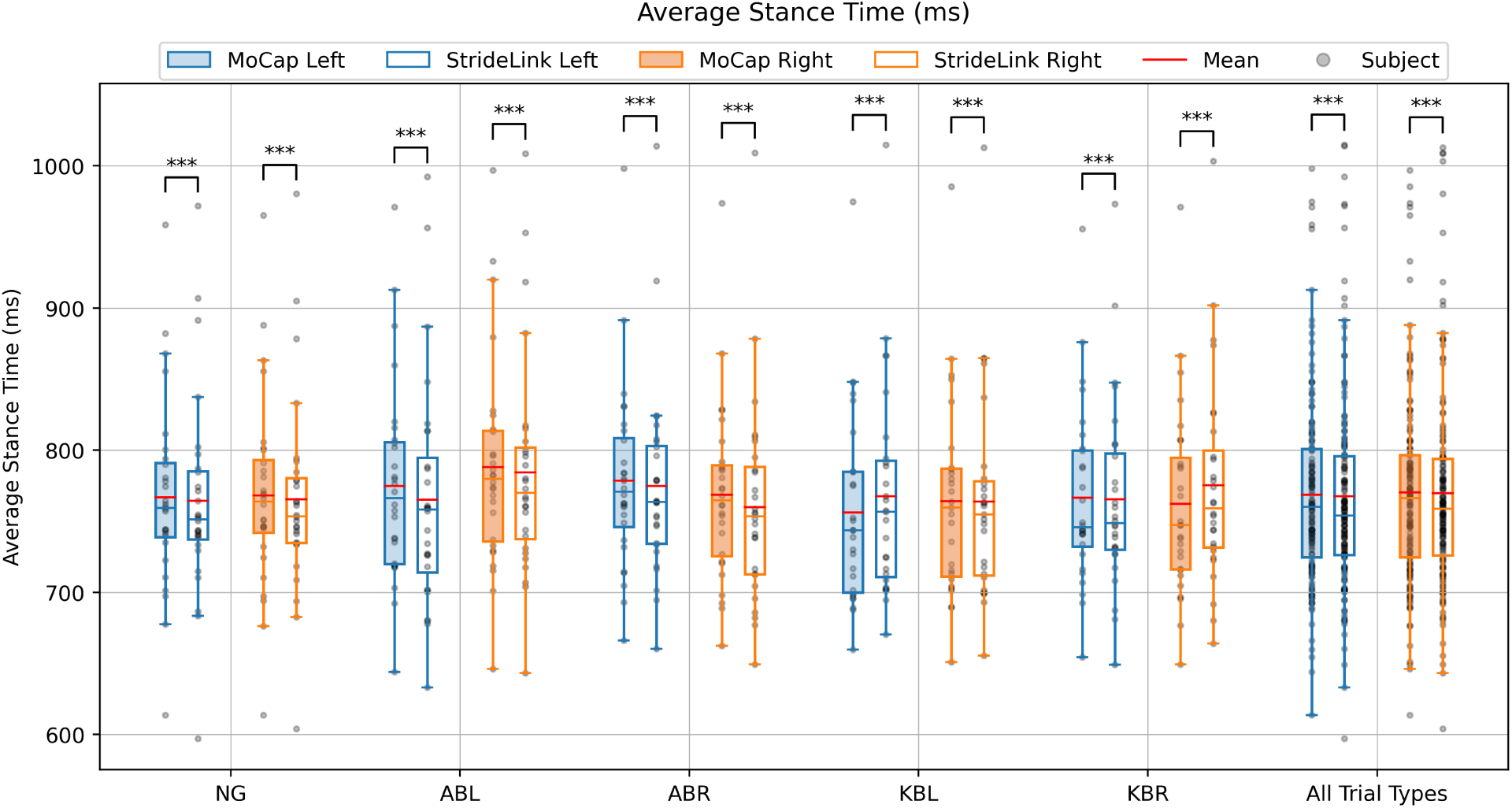
Boxplots of average stance time measured with motion capture and the StrideLink device. Motion capture boxplots are shaded and device boxplots are not. Left lower limb metrics are shown in blue, and right lower limb metrics are shown in orange. Mean values are shown as a red line within each boxplot. Individual subject mean data points are shown as points. Trial type abbreviations are as follows: NG - normal gait, ABL - ankle boot left, ABR - ankle boot right, KBL - knee brace left, KBR - knee brace right

## 4. Discussion

Stride time measured with the StrideLink device was significantly equivalent to stride time measured with motion capture. The StrideLink device underestimated the motion capture measurements by an absolute error of approximately -3.75 ms on average, which corresponded to a relative error of less than 1%. This error is lower than the approximate 5% error values reported by Fusca *et al.* [29] when comparing a single waistworn IMU to motion capture, and Agostini *et al.* [30] when comparing IMUs worn on the shank and toe of each foot to motion capture. The lower error percentage reported in this study may be attributed to both the placement of StrideLink sensors (worn on the top of the foot) and the processing algorithms that the system utilizes, resulting in a more accurate capture of stride time. In a broader review, Kobsar *et al.* investigates the accuracy of stride time calculations across IMU placements and processing methods and finds both foot and lumbar placement to show excellent validity and reliability which may further suggest the demonstrated improved accuracy may be more likely attributed to the processing methods rather than placement alone [19]. Notably, these studies did not assess the accuracy of the system under abnormal walking conditions.

Cadence results also showed good agreement between the StrideLink device and motion capture. The device overestimated cadence by approximately 0.85 steps/minute on average, producing mean relative errors below 1%. The consistently overestimated cadence of the device may be a result of the slight difference between calculation methods of the device and motion capture. The StrideLink device’s calculation uses the average stride time to estimate the cadence, whereas the motion capture uses the total number of left and right steps in the steady state portion of the trial. Calculating the number of individual steps results in a single step difference as the motion capture data is cropped to include the same number of left and right strides. Additionally, there may be a statistical pattern to the gait cycles being dropped by the device, resulting in a systematic underestimation of gait cycle times and an increase in apparent cadence. Nonetheless, the relative error in cadence is lower than the approximately 5% values reported by Fusca *et al.* [29], Agostini *et al.* [30] and Yeo *et al.* [17] all of which assessed healthy walking patterns in young adults utilizing various IMU placement methods.

Double support time results did show some equivalence between the gait measurement device and motion capture measurements, but there still appeared to be errors in agreement between the measurement methods. Double support time relative errors were high (above 5%) for a large number of subjects suggesting that individual outlier subjects were not the reason for non-equivalence. A consequence of the calculation method is that double support time depends on the leading foot to determine the 1st and 2nd double support phases of a gait cycle. As the StrideLink device removes some gait cycles during analysis, the leading foot may have alternated between periods of analysis. This would have changed the 1st and 2nd double support phases for each gait cycle and potentially impacted the gait metric results. Additionally, if the device’s algorithm did not remove the first double support phase in a region of interest, the average double support time may have been skewed. In future work, the accuracy of double support time measurements should be investigated under standardized protocols, especially seeing as studies conducted by Berner *et al.* [8] who used a multi-sensor lower limb setup to compare subjects with and without gait impairments, and Kvist *et al.* [31] which integrated waist and foot-worn IMU sensor processing methods, have reported similarly high double support time relative errors.

While swing time results were equivalent across all trial types, LCC agreement was lacking. The increased error in swing times for ankle boot trials is consistent with the findings of a study conducted by Jocham *et al.* [11] that investigated the influence of shoe motion on the agreement between IMU and motion capture measurements. The effect of motion capture marker placement on the obtained measurement was found to be as large or larger than than discrepancies between motion capture and IMU measurements. The placement of IMUs for the present study was based on findings from Niswander *et al.* [18] and agrees with recent findings by Kvist*et al.* showing high accuracy at foot-worn locations compared to shank or lumbar sensors when gait patterns are altered [31]. This offers further support for use of foot-worn IMUs, but does suggest future work to be conducted to find the optimal placement for optical markers on non-standard footwear.

Stance time measurements taken with the StrideLink device aligned very well with motion capture measurements. Mean absolute error of stance time was less than 20 ms for each foot and trial type, typically resulting in mean relative error percentages of less than 1%. These mean relative error values are comparable to those reported by Laidig *et al.* [20] who similarly investigated the accuracy of foot-worn IMU sensors under abnormal conditions. No clear biases were noted in the stance time absolute error. Pearson correlations were high to very high, but Lin’s correlation coefficient values were often poor. Further tuning of the device’s algorithm may be warranted to improve LCC values.

In general, the StrideLink device maintained similar measurement accuracy and agreement with motion capture measurements under normal and abnormal gait conditions. These results are more promising than the study findings of Benoussaad *et al.* [21], Chalmers *et al.* [25], and Schwameder *et al.* [24], all of which indicate that IMU accuracy decreases when analyzing simulated gait deviations including obstacles and shoe-height changes to induce a limp. The agreement between IMU and motion capture measurements is furthermore consistent with the more recent findings of Jocham *et al.* [11] and Laidig *et al.* [20]. Compared to other algorithms and methods for gait detection using IMUs, StrideLink offers a novel calibration-free approach that uniquely applies changes in acceleration to detect gait events meeting the need for an easy-to-use, highly accurate sensor that can be applied to everyday clinical settings.

The method utilized to produce abnormal gait (knee and ankle braces) did produce gait alterations, particularly resulting in increased variability across subjects, especially for the knee brace.

Of note, the abnormal gait shows some utility in validating gait analysis procedures; in the average swing time (Figure 7), a greater difference between StrideLink and the Vicon results can be seen specifically for the limbs wearing the knee brace, resulting in an underestimation of swing time by about 10 ms. This difference was only obtained in that particular case of altered gait, and would have been missed had only normal gait been measured.

## 5. Conclusion

In summary, this study demonstrated that the majority of gait parameters determined by StrideLink’s new IMU-based gait measurement device were equivalent to and correlated well with motion capture measurements. Particularly high degrees of agreement were achieved for the gait metrics of stride time, cadence, and stance time. The StrideLink device additionally maintained sufficient measurement accuracy and agreement with the motion capture measurements under conditions of normal gait, and abnormal gait induced by an ankle boot or knee brace. The use of abnormal gait as part of the validation provides greater confidence that the StrideLink device can be used with users possessing or acquiring abnormal gait characteristics, and this work additionally provides some evidence of the utility of our particular method of inducing abnormal gait for the purpose of validating gait measurement systems.

## Data Availability

All data produced in the present study are available upon reasonable request to the authors

## CRediT Authorship Contribution Statement

**Aaron Henry:** Investigation, Writing – original draft, Methodology, Formal analysis, Visualization, Conceptualization. **Carson Benner:** Investigation, Writing – review & editing. **Andrew Robbins:** Writing – review & editing, Methodology, Conceptualization, Supervision. **Cassandra McIltrot:** Writing – review & editing, Methodology, Conceptualization.

## Appendix A. Supporting Information

Supplementary data associated with this article can be found in the online version.

## Acknowledgements

The authors wish to thank StrideLink, Inc. for providing funding for this work and all subjects for participating in the study.

---

**Supplemental Table 1:** Pearson Correlation Coefficients for each trial type and gait metric. Green cells indicate r > 0.9, blue cells indicate r > 0.8, and yellow cells indicate r > 0.7.

| Pearson Correlation Coefficients (PCCs) |  |  |  |  |  |  |  |  |
| --- | --- | --- | --- | --- | --- | --- | --- | --- |
| Metric | Normal Gait | Ankle Boot Left | Ankle Boot Right | Knee Brace Left | Knee Brace Right | All Left | All Right | All |
| Average Stride Time Left (ms) | 1.000 | 0.999 | 1.000 | 0.999 | 0.999 | 0.999 |  |  |
| Average Stride Time Right (ms) | 0.999 | 1.000 | 0.999 | 1.000 | 0.999 |  | 0.999 |  |
| Average Cadence (steps/min) | 0.999 | 0.999 | 0.999 | 0.999 | 0.999 |  |  | 0.999 |
| Average Double Support Time (ms) | 0.946 | 0.957 | 0.953 | 0.964 | 0.960 |  |  | 0.940 |
| Average Double Support Pct (%) | 0.869 | 0.882 | 0.880 | 0.910 | 0.897 |  |  | 0.855 |
| Average Swing Time Left (ms) | 0.837 | 0.728 | 0.856 | 0.912 | 0.891 | 0.826 |  |  |
| Average Swing Left Pct (%) | 0.841 | 0.829 | 0.848 | 0.927 | 0.813 | 0.807 |  |  |
| Average Swing Time Right (ms) | 0.862 | 0.915 | 0.742 | 0.913 | 0.889 |  | 0.834 |  |
| Average Swing Right Pct (%) | 0.849 | 0.922 | 0.833 | 0.851 | 0.924 |  | 0.826 |  |
| Average Stance Time Left (ms) | 0.988 | 0.980 | 0.988 | 0.991 | 0.990 | 0.982 |  |  |
| Average Stance Left Pct (%) | 0.842 | 0.831 | 0.846 | 0.929 | 0.813 | 0.808 |  |  |
| Average Stance Time Right (ms) | 0.987 | 0.995 | 0.982 | 0.991 | 0.991 |  | 0.984 |  |
| Average Stance Right Pct (%) | 0.848 | 0.924 | 0.835 | 0.850 | 0.924 |  | 0.827 |  |

**Supplemental Table 2:** Lin’s Correlation Coefficients for each trial type and gait metrics. Green cells indicate r > 0.99 and are interpreted as almost perfect agreement, blue cells indicate r > 0.95 and substantial agreement, yellow cells indicate r > 0.90 and moderate agreement, orange cells indicate r < 0.9 and indicate poor agreement, red cells indicate r < 0.8 and poor agreement.

| Lin's Correlation Coefficients (LCCs) |  |  |  |  |  |  |  |  |
| --- | --- | --- | --- | --- | --- | --- | --- | --- |
| Metric | Normal Gait | Ankle Boot Left | Ankle Boot Right | Knee Brace Left | Knee Brace Right | All Left | All Right | All |
| Average Stride Time Left (ms) | 0.998 | 0.999 | 0.999 | 0.999 | 0.999 | 0.999 |  |  |
| Average Stride Time Right (ms) | 0.998 | 0.999 | 0.998 | 0.999 | 0.998 |  | 0.999 |  |
| Average Cadence (steps/min) | 0.990 | 0.992 | 0.991 | 0.992 | 0.991 |  |  | 0.991 |
| Average Double Support Time (ms) | 0.900 | 0.926 | 0.918 | 0.864 | 0.846 |  |  | 0.891 |
| Average Double Support Pct (%) | 0.768 | 0.818 | 0.818 | 0.683 | 0.671 |  |  | 0.749 |
| Average Swing Time Left (ms) | 0.827 | 0.708 | 0.855 | 0.788 | 0.879 | 0.821 |  |  |
| Average Swing Left Pct (%) | 0.820 | 0.666 | 0.830 | 0.712 | 0.792 | 0.759 |  |  |
| Average Swing Time Right (ms) | 0.858 | 0.911 | 0.720 | 0.901 | 0.697 |  | 0.823 |  |
| Average Swing Right Pct (%) | 0.837 | 0.909 | 0.686 | 0.833 | 0.709 |  | 0.788 |  |
| Average Stance Time Left (ms) | 0.986 | 0.962 | 0.984 | 0.975 | 0.988 | 0.978 |  |  |
| Average Stance Left Pct (%) | 0.822 | 0.663 | 0.826 | 0.723 | 0.792 | 0.761 |  |  |
| Average Stance Time Right (ms) | 0.986 | 0.993 | 0.965 | 0.990 | 0.971 |  | 0.982 |  |
| Average Stance Right Pct (%) | 0.838 | 0.910 | 0.682 | 0.834 | 0.718 |  | 0.791 |  |

